# Epidemiological impact and cost-effectiveness of expanding formal PrEP provision to PrEP-eligible MSM expressing PrEP-intention in the Netherlands

**DOI:** 10.1101/2023.06.19.23291600

**Authors:** Haoyi Wang, Stephanie Popping, David van de Vijver, Kai. J. Jonas

## Abstract

**Objective:** Reimbursed pre-exposure prophylaxis (PrEP) access in the Netherlands has reached its maximum capacity with a waiting-list of 3,000 men-who-have-sex-with-men (MSM) and 19,500 PrEP-eligible/intending MSM. This study models the epidemiological impact and cost-effectiveness of expanding PrEP provision to waiting-list and PrEP-eligible/intending MSM in the Netherlands, given the imminent national evaluation of the current PrEP programme in July 2023.

**Methods:** We calibrated a deterministic transmission model of HIV among MSM. Expanded PrEP provision was seeded in 01/2022, to achieve the coverage (varied at 25%,50%,75%, and 100%) for the waiting-list (n=3,000), and PrEP-eligible/intending group (n=19,500) by 01/2024. The epidemiological impact was modelled until 2030, while cost-effectiveness and the budget impact were calculated from a payer’s perspective over 40-years, and five-years, starting from 2022, respectively.

**Results:** Expanding PrEP provision leads to further reductions in HIV infections among MSM. Covering waiting-list and PrEP-eligible/intending MSM can avert a total of 11 (2.5%) up to 192 (45.1%) new infections by 2030. Expanding PrEP provision to over-75% of PrEP-eligible/intending MSM offers the possibility of ending the HIV epidemic by 2030. However, achieving this milestone comes with significant costs, with an incremental-cost-effectiveness-ratio of €164,100 per quality-adjusted-life-year and short-term costs of €1,074 million over five-years.

**Conclusions:** This study provides timely evidence for the upcoming national evaluation of the PrEP program in the Netherlands, supporting its continuation and further expansion. While expanding PrEP provision holds promise for HIV elimination, given the associated significant costs, careful consideration is crucial to balance efforts to end the HIV epidemic and the available resources.

## Introduction

HIV pre-exposure prophylaxis (PrEP) is formally available in the Netherlands since 2019 [1], primarily targeting men-who-have-sex-with-men (MSM) [2]. However, the current formal PrEP provision through public health services is reaching its maximum capacity of 8,500 slots [3]. Via this formal access pathway, PrEP can be provided to PrEP-eligible MSM with a very low co-payment due to a national pilot programme subsidy from the Dutch Minister of Health for a period of five years from 2019 onwards [4]. Yet, the current formal PrEP provision pathway has almost reached maximum capacity [3], with approximately 3,000 MSM on waiting-list (hereinafter waiting-list MSM) [3, 5].

Yet, this number likely underestimates the actual demand, as it does not account for other pathways of PrEP use, such as through general practitioners (GPs) or online pharmacies outside the formal national registry system, where the national subsidy and the discounted co-payment do not apply [6]. Consequently, there is a consensus in the Netherlands that there are more MSM that meet the Dutch PrEP eligibility criteria [1], are willing to use PrEP (hereinafter PrEP-eligible/intending MSM) and are waiting for formal PrEP access [4, 6]. Therefore, the size of the PrEP-eligible/intending MSM population should be estimated to understand the unmet needs of PrEP for better PrEP provision and uptake.

More importantly, the imminent national evaluation of the current PrEP pilot programme, scheduled by and starting at the end of July 2023, will determine the future of the national PrEP programme and the national subsidy for PrEP uptake provided by the Dutch Minister of Health from 2024 onwards. Given the Dutch commitment to ending the HIV epidemics by 2030 [7], the substantial costs of HIV care and the importance of allocating healthcare budgets wisely, it is vital to consider both epidemiological and health economic evidence to inform decision-making for the future HIV prevention strategies in the Netherlands, especially under a context of a declining epidemic [8].

Therefore, to contribute to this evaluation process and provide timely evidence, this study models the epidemiological impact and cost-effectiveness of continuing and expanding formal PrEP provision to waiting-list MSM and PrEP-eligible/intending MSM in the Netherlands.

## Methods

### Estimating the number of PrEP-eligible/intending MSM

To estimate the number of PrEP-eligible/intending MSM living in the Netherlands, we utilised the Dutch subsample of the European-MSM-Internet-Survey (EMIS-2017) [6, 9]. Following the Dutch PrEP eligibility guideline [1], MSM who have had 1) condomless anal intercourses with partner(s) with unknown or seropositive HIV status, 2) syphilis, rectal chlamydia or gonorrhoea and/or 3) a post-exposure prophylaxis prescription in the preceding six months are considered to be PrEP-eligible [1]. Expressing intention for PrEP use was defined as being quite- and very-likely to use PrEP when PrEP is available and affordable [9, 10].

We estimated that approximately 35% of HIV-negative MSM were PrEP-eligible [1, 6]. Among those MSM, 45% expressed intentions for PrEP use [6]. Given the estimated size of the HIV-negative MSM population in the Netherlands [11], this results in 28,000 PrEP-eligible/intending MSM who would also benefit from PrEP, and a striking 19,500 PrEP-eligible/intending MSM with unmet PrEP needs.

### Mathematical transmission model assumptions and calibration

In this study, we utilised an existing transmission model of HIV among MSM [12], to represent the HIV epidemic among MSM aged 15 years and older in the Netherlands. Schematic representations of the model, the equations, and the assortative mixing matrix by sexual risk groups were previously published (Supplement pp1–6) [12]. In short, our model stratifies disease progression into the acute stage, three chronic stages (CD4>500 cells/mm^ 3, CD4 count 350–500 cells/mm^ 3, and CD4 count 200–349 cells/mm^ 3) and one AIDS stage (CD4 <200 cells/mm^ 3). Our model included four different sexual risk groups with different levels of sexual activity based on the annual number of new sexual partners [12, 13].

The model was calibrated to the historical Dutch HIV epidemic among MSM from 2017 to 2021 [8]. The key model parameters were outlined and further specified in Supplementary Table S1 through S2. We accepted 117 of 500,000 simulations that best matched the Dutch HIV epidemic among MSM [8]. The calibrated model included in this study was previously published [13]. All results are reported as the median of the accepted simulations.

The base-case scenario simulated the current Dutch formal PrEP provision and the HIV care continuum to represent the current and future HIV epidemic given the current oral PrEP services and serve as counterfactual estimates when evaluating the expanded PrEP provision scenarios. The base-case model characteristics in terms of HIV incidence, the proportion of the population and estimated HIV incidence in each sexual activity group in 2022 and the proportion of antiretroviral treatment (ART) use among MSM with HIV were previously published [13]. The expanded PrEP provision was seeded in January 2022, allowing a two-year scaling-up period, to achieve the coverage (varied at 25%,50%,75%, and 100%) for the waiting-list (3,000 MSM), and PrEP-eligible/intending group (19,500 MSM) by January 2024 (Figure S1).

### Epidemiological impact

Committing to ending the HIV epidemic by 2030 [7], we modelled the epidemiological impact of all scenarios until 2030 based on new HIV infections over time, and the cumulative HIV infections (percentage) averted in comparison with the base-case scenario.

### Cost-effectiveness and budget impact

The Consolidated Health Economic Evaluation Reporting Standards (CHEERS) checklist can be found in Supplementary S2 File. Cost-effectiveness was calculated from a payer’s perspective with a lifetime horizon of 40 years from 2022 onwards. In our deterministic model, each compartment was assigned a quality-adjusted life year (QALY) score and a cost. Costs and QALY were discounted at 3% per year. Incremental cost-e□ectiveness ratios (ICER) were determined, using a Dutch willingness-to-pay threshold set at <€20,000, 20,001-50,000 and 50,001-80,000 per QALY [14]. Healthcare resource unit prices were based on recommended prices by the Dutch Healthcare Institute and the Dutch Healthcare Authority [15], while the healthcare utilisations were based on clinical data from the Dutch MSM population, stratified by the diagnosed stages [16] (Supplementary Table S3 through S5 for detailed utility weighting and costs). To better inform decision-making, we additionally calculated the short-term budget impact of all scenarios over five years [17].

For the scenario with the most optimal epidemiological impact, we performed a univariate sensitivity analysis of the cost-effectiveness of the expanded PrEP provision towards PrEP-eligible/intending MSM compared with the current PrEP provision (base-case scenario) in which we varied five key input variables. QALY discounting was varied between 0–3%, and cost discounting between 0–4·5%. The yearly cost of ART was varied with 15%. The monthly cost of PrEP was decreased to seven euros per month, given the current co-payment of oral PrEP in the Netherlands [6]. Lastly, the number of screenings for sexually STIs (including HIV, syphilis, hepatitis C and other bacterial STIs) among susceptible MSM who are not on PrEP ranged to once per two years.

## Results

### Epidemiological impact of expanding formal PrEP provision

Our model showed that covering 100%, 75%, 50%, or 25% of the waiting-list MSM by January 2024 resulted in a total number of 39 (9.1%), 30 (7.0%), 20 (4.8%), or 11 (2.5%) new HIV infections averted by 2030, respectively (Figure 1). Most noteworthy, further expanding PrEP provision towards 100% or 75% of PrEP-eligible/intending MSM provides the possibility to end the HIV epidemic in 2030 (zero new infection in 2030). By 2030, a total number of 58 (13.5%) up to 192 (45.1%) can be averted related to the coverage levels of PrEP to PrEP-eligible/intending MSM (Figure 2).

**Figure 1.**
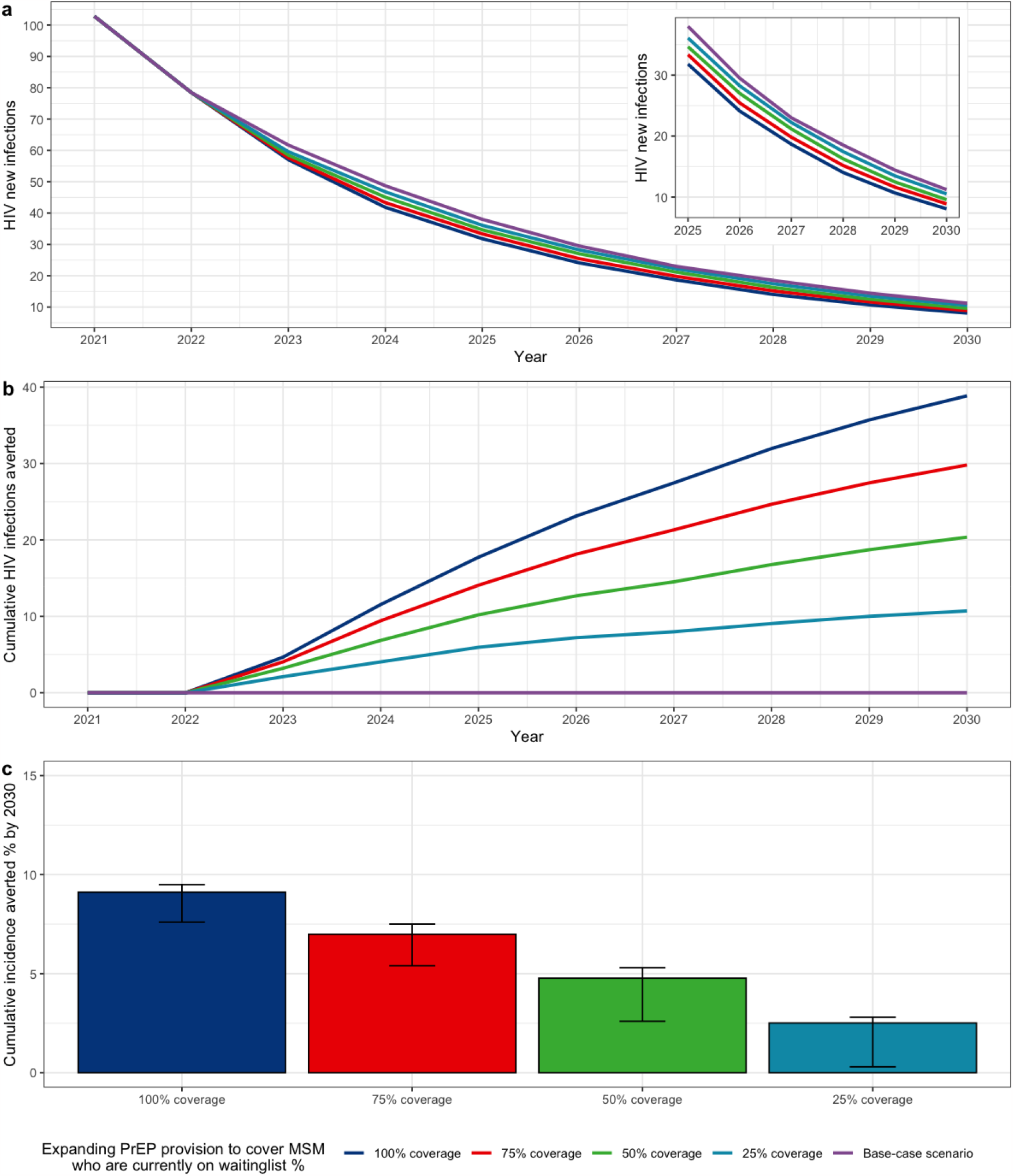
Projected a) HIV new infections, b) cumulative HIV new infections averted, and c) cumulative HIV new infections averted percentage by 2030 by further expanding PrEP provision to MSM who are currently on the waiting list with formal PrEP access, the Netherlands, 2021-2030. Note: Lines show median across 117 fits. Bars and error bars show median and interquartile range across 117 fits.

**Figure 2.**
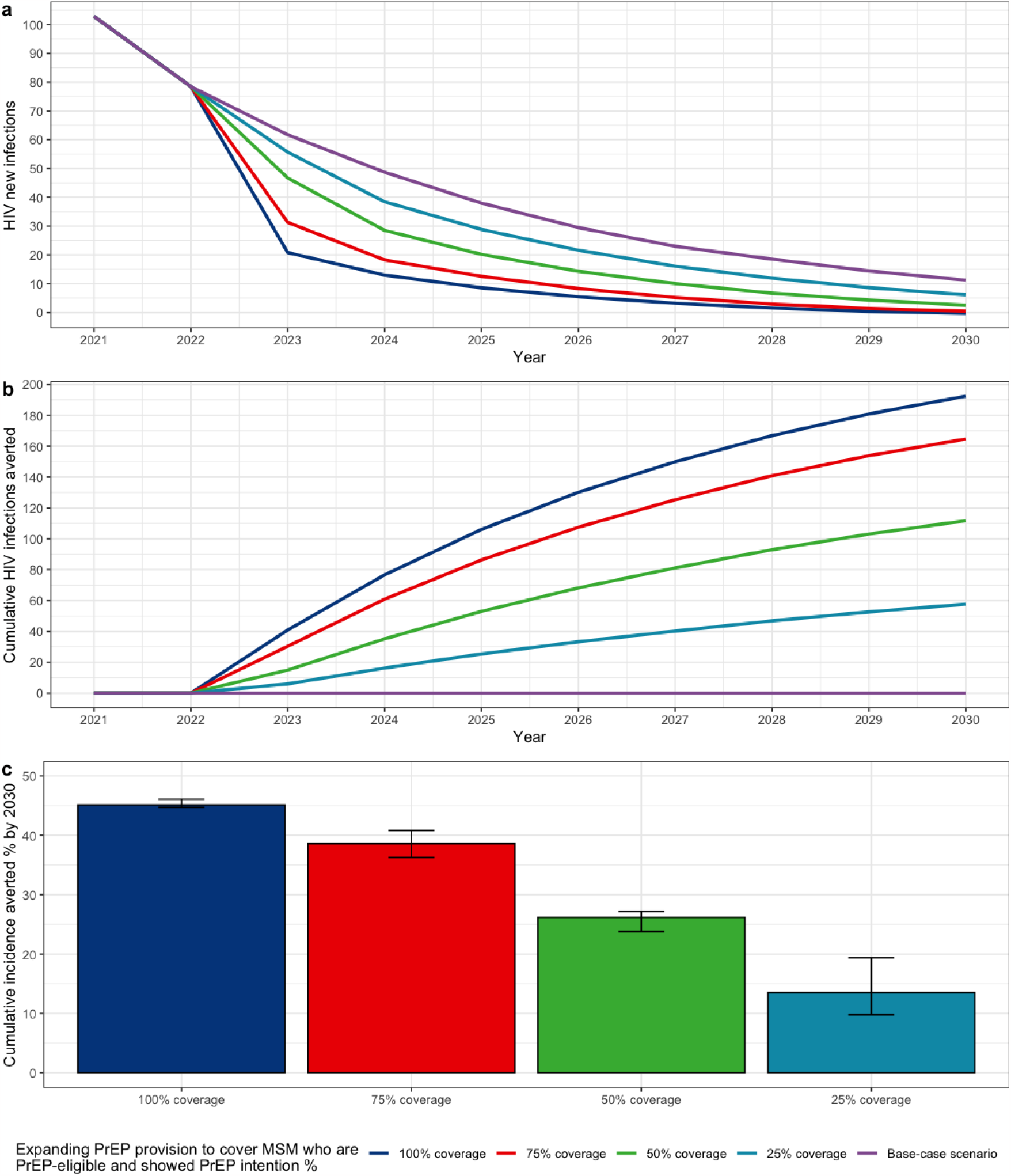
Projected a) HIV new infections, b) cumulative HIV new infections averted, and c) cumulative HIV new infections averted percentage by 2030 by further expanding PrEP provision to MSM who are PrEP-eligible and express intention to use PrEP with formal PrEP access, the Netherlands, 2021-2030. Note: Lines show median across 117 fits. Bars and error bars show median and interquartile range across 117 fits.

### Cost-effectiveness of expanding formal PrEP provision

Our model showed that none of the scenarios was cost-saving (Figure 3). Specifically, for the waiting-list scenarios, achieving 25% coverage by January 2024 was forecasted to be cost-effective with an ICER of €7,600 per QALY. However, achieving 100% coverage resulted in an ICER of €43,000 per QALY. For the PrEP-eligible/intending scenarios, achieving 50% coverage was forecasted to be the most cost-effective with an ICER of €38,600. Although achieving 75% coverage was forecasted in zero new infections in 2030, this scenario resulted in an ICER of €164,100 per QALY and therefore not cost-effective. Our budget impact analysis showed that reaching zero new HIV infections by 2030 would cost €1,074 million over five years (see Table 1 for other scenarios).

**Figure 3.**
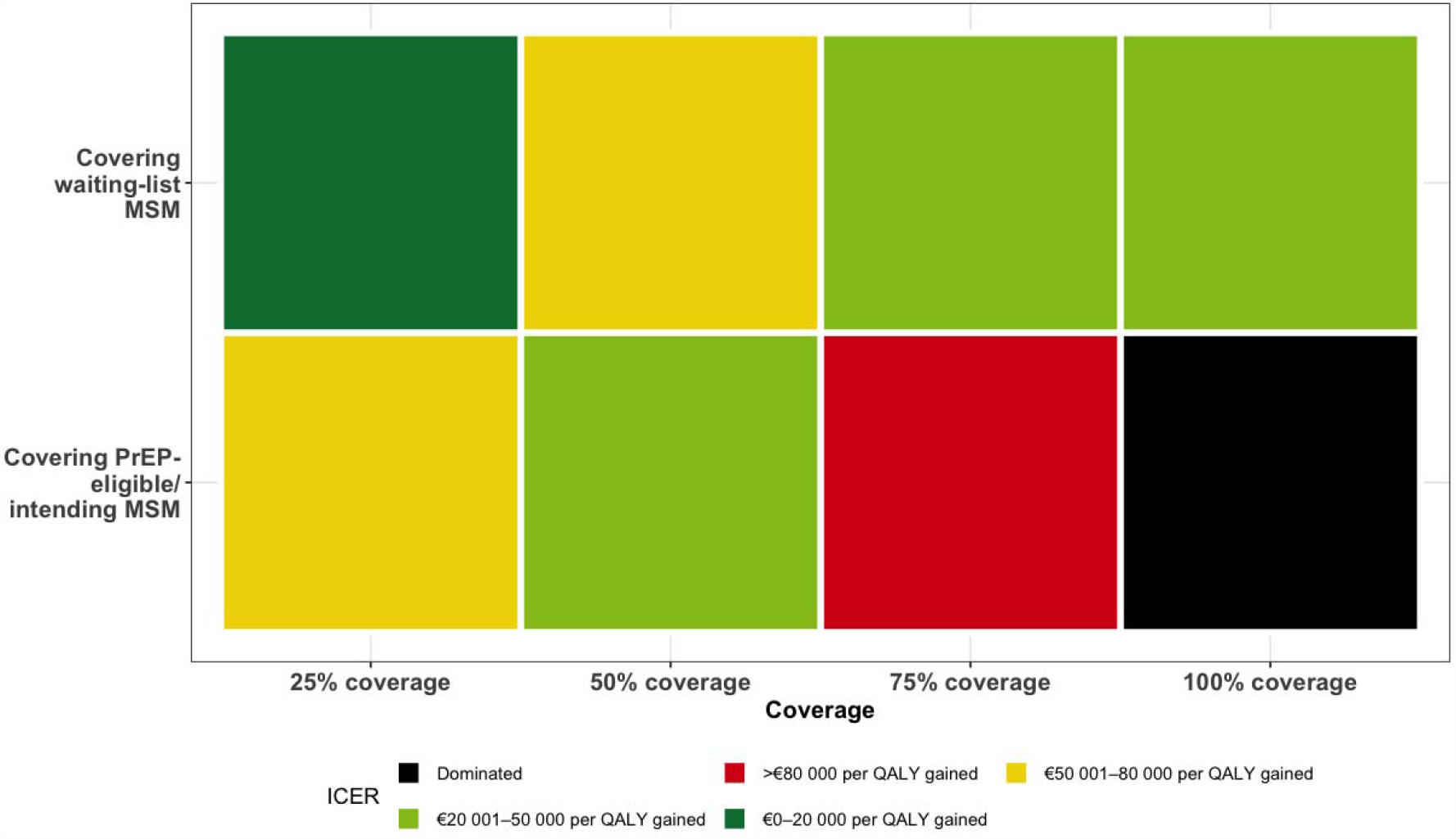
Cost-effectiveness analysis in incremental cost-effectiveness ratio (ICER) of expanding PrEP provision over the different scenarios, the Netherlands, 2022-2062. Note: Cost-effectiveness ratios are provided over the different scenarios using a lifetime horizon of 40 years. Expanding PrEP provision is started in January 2022. The waiting-list group contains 3.000 MSM and the eligible/intending group contains 19.500 MSM. Coverage is reached by January 2024. Abbreviations: ACER: ICER: incremental cost-effectiveness ratio; MSM: men who have sex with men; PrEP: pre-exposure prophylaxis; QALY: Quality Adjusted Life Years. Dominated: When the compared strategy has equal or less QALYs compared with the previous less costly scenario.

**Table 1.**
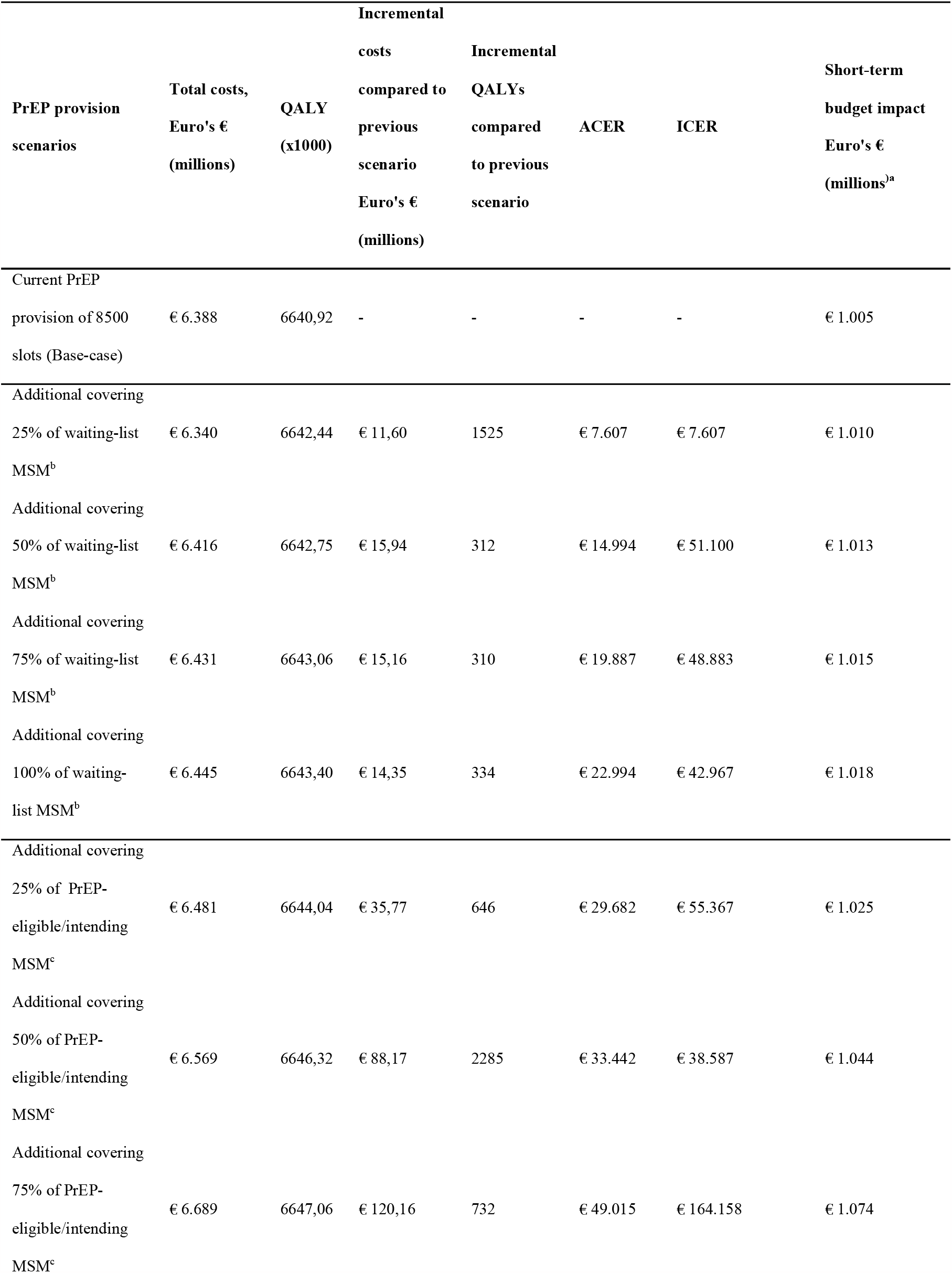
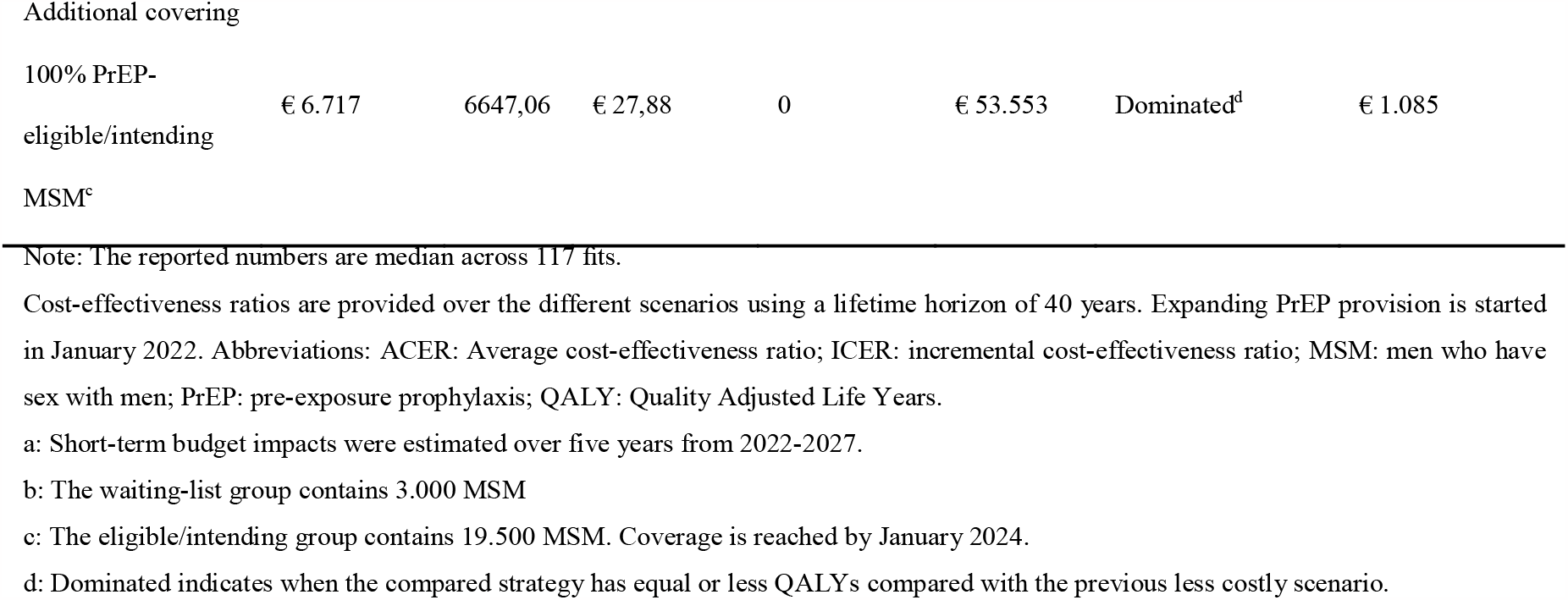
Cost-effectiveness of expanding PrEP provision over the different scenarios, the Netherlands, 2022-2062.

We performed the sensitivity analysis on the scenario covering 75% of PrEP-eligible/intending MSM by January 2024 compared to the base-case scenario. Our one-way sensitivity analyses highlighted that our cost-e□ectiveness analysis is most sensitive to the yearly cost discounting rate. Improving the PrEP provision to cover 75% of PrEP-eligible/intending MSM by January 2024 can be considered 25% more cost-effective if 4.5% of yearly cost discounting was assumed, and 75% less cost-effective if 0% of yearly cost discounting was assumed. In addition, our cost-e□ectiveness analysis is also sensitive to the monthly price of oral PrEP that improving PrEP provision to cover 75% of PrEP-eligible MSM with PrEP-intention by January 2024 can be considered 40% more cost-effective if a seven-euro monthly cost was assumed. More details can be found in Figure S2.

## Discussion

Our study aimed to compare the epidemiological impact and cost-effectiveness of expanding formal PrEP provision in comparison to the current pilot programme. We consider our results as timely for the upcoming national evaluation of the current PrEP programme in the Netherlands, and our results would also remain relevant for other settings with a declining HIV epidemic like the one in the Netherlands.

We found that expanding formal PrEP provision leads to further reductions in HIV infections among MSM with the possibility of HIV elimination by 2030. Our findings thus commit to the goal to end the HIV epidemic by 2030 [7, 18]. Specifically, we forecasted that providing PrEP to over 75% of the PrEP-eligible/intending MSM would lead to zero new infections by 2030. These results corroborate other studies emphasizing the substantial impact of reducing HIV incidence by increasing PrEP provision, access, and usage [13, 19]. However, the current PrEP uptake in the Netherlands faces challenges due to limited provision capacity within public health services, and many GPs perceive PrEP provision as beyond their scope [20]. Consequently, the real-life PrEP uptake may fall short of achieving the optimal PrEP coverage scenarios demonstrated in this analysis (over 75% PrEP eligible/intending MSM). To address this, a centralised PrEP registry should be established, and a de-centralisation of PrEP provisions to encourage other medical doctors, including GPs, deems necessary and should be implemented. These approaches align with practices in other European countries like France [10] and Belgium [21], and the elimination programmes of other STIs, such as hepatitis C virus [22]. In turn, the formal provision and uptake of PrEP would be enhanced to support ending the HIV epidemic.

However, despite emphasising the potential major epidemiological benefits, our findings also highlight the significant costs associated with achieving zero new HIV infections by 2030 through PrEP expansion alone considering the current prices. Such a high cost was expected, given that zero new HIV infections always moves in an expected U-shape in terms of the costs of other HIV-related services/programmes [23]. Alternatively, expanding PrEP to 25% of the waiting-list MSM or 50% of PrEP-eligible/intending MSM could be considered a cost-effective strategy. Yet, these scenarios were not predicted to reach zero new HIV infections by 2030. Therefore, a trade-off between the desired epidemiological impact and the associated costs necessitates careful consideration during the upcoming evaluation. When deciding to pursue the goal of zero new HIV infections by 2030, a high-level Dutch political and societal commitment [7] would be needed. Our sensitivity analysis suggested that reducing the current price of PrEP would likely make the path to zero new HIV infections by 2030 more affordable.

One major strength of this study was the use of the well-described Dutch data on the MSMHIV epidemic [8], the use of the most comprehensive MSM sexual behavioural data in the Netherlands from the Dutch subsample of EMIS-2017 [6], and the use of the well-validated and calibrated model which described and predicted the HIV epidemic among MSM [12, 13]. In turn, these allowed us to make accurate epidemic predictions. Another major strength of this study was the use of the healthcare utilisation stratified by the diagnosed stages for HIV treatment and its associated local costs. Consequently, our model did not overlook the potential impact of the late and advanced presentation of HIV diagnoses, which are not uncommon in the Netherlands [8].

This analysis has several limitations. Firstly, our model assumed a daily-use scenario for PrEP. Given the prevalent event-driven PrEP use in the Netherlands [24, 25], our estimations may not be generalisable to the event-driven PrEP-using population. Future evaluation should consider this subpopulation when the accurate size of event-driven PrEP users is available. In addition, our model assumed only MSM with higher sexual risk would use PrEP [13], while this may not be reflected in real-life contexts and practices. In turn, our projected epidemiological impacts and cost-effectiveness may be overestimated.

## Conclusions

Our study provides timely evidence for the upcoming national PrEP evaluation programme to support the further expansion of PrEP provision in the Netherlands. Our long-term cost-effectiveness and short-term budget impact analysis offer valuable insights for decision-making, considering the epidemiological impacts and costs of different levels of PrEP expansion. We emphasize the potential benefits of expanding the current PrEP programme to reach more eligible individuals through a centralised PrEP registry and de-centralised provision. However, expanding the programme to eliminate HIV comes with significant costs. Therefore, careful societal and political considerations are needed to strike a balance between efforts to end the HIV epidemic and the available resources.

## Data Availability

All data produced in the present study are available upon reasonable request to the authors.

## Notes

**Conflict of interest:** HW, DvdV, and KJJ report grants from ViiV Healthcare outside the submitted work. DvdV and KJJ also report grants from Gilead Sciences outside the submitted work.

### Competing Interest Statement

HW, DvdV, and KJJ report grants from ViiV Healthcare outside the submitted work. DvdV and KJJ also report grants from Gilead Sciences outside the submitted work.

### Funding Statement

This study did not receive any funding.

### Author Declarations

Data retrieved from European MSM Internet Survey 2017 was presented in an aggregated and anonymised manner. Ethical approval and informed consent were obtained from the Observational Research Ethics Committee at the London School of Hygiene & Tropical Medicine (review reference 14421/RR/8805). For other data used in this study, ethical approval and informed consent were waived. The study was conducted in line with the Consolidated Health Economic Evaluation Reporting Standards guidelines.

