## Supplemental File S1 for "Epidemiological impact and cost-effectiveness of expanding formal PrEP provision to PrEP-eligible MSM expressing PrEP-intention in the Netherlands"

**Supplementary S1 File**

Haoyi WANG^1,2*^, Stephanie POPPING^2,3^, David VAN DE VIJVER^2^, Kai. J. JONAS ^1^

^1^Department of Work and Social Psychology, Maastricht University, Maastricht, The Netherlands

^2^Viroscience department, Erasmus Medical Centre, Rotterdam, The Netherlands

^3^Department of Medical Microbiology and Infectious Diseases, Erasmus Medical Centre, Rotterdam, The Netherlands

* Correspondence to: Haoyi Wang

### Model calibration

We calibrated our model to the historical epidemic based on: the estimated size of the MSM population, the number of oral PrEP users, the number of MSM diagnosed with HIV, the estimated number of MSM living with HIV, the yearly number of new HIV diagnoses including the proportion diagnosed in a late stage (CD4<350 cells/mm^3) and advanced stage of infection (CD4<200 cells/mm^3), and the proportion of HIV diagnosed MSM receiving antiretroviral treatment, variables that were used for model calibration can be found in Table S1. Key model parameters can be found in Table S2. Latin Hypercube sampling was used to calibrate the model.

**Table S1 Variables used to calibrate and accept simulation using Latin Hypercube sampling techniques.**

| **Parameter used for calibration** | | **Data in real world** | **Values accepted in calibration range** | **Source** |
| --- | --- | --- | --- | --- |
| MSM population 2018-2020 | | 131515 | 130000 - 200000 | [1] |
| The number of oral PrEP users 2022 | | 8500 | 7000 - 11000 | [2, 3] |
| Number of MSM diagnosed with HIV 2020 | | 13876 | 12500 - 15000 | [4] |
| The estimated number of MSM living with HIV 2020 | | 14500 | 12000 - 16000 | [4] |
| Number of new diagnosis amiong MSM | |  |  | [4] |
|  | 2018 | 446 | 300 - 600 |  |
|  | 2019 | 340 | 200 - 500 |  |
|  | 2020 | 257 | 100 - 400 |  |
| Proportion late diagnosis | | 42% | 37% - 47% | [4] |
| Proportion advanced diagnosis | | 22% | 17% - 27% | [4] |
| The proportion of HIV diagnosed MSM receiving ART | |  |  | [4] |
|  | 2020 | 92% | 87% - 98% |  |
|  | 2021 | 96% | 91% - 100% |  |

^Note: A total of 117 simulation were accepted (out of 50000 simulations run).^

Table S2. Key model parameters of PrEP for HIV prevention in the Netherlands

| **Model parameters** | | | **Estimate or range^a^/IQR** | **Reference** |
| --- | --- | --- | --- | --- |
| Duration of disease stages | | |  |  |
|  | Acute stage | | 10–16 weeks | [5] |
|  | CD4+ T-cell count 350–500 cells/µL | | 2.9–3.1 years | [6] |
|  | CD4+ T-cell count 200–349 cells/µL | | 3.6–3.9 years | [6] |
|  | CD4+ T-cell count < 200 cells/µL | | 13–25 months | [6] |
| Infectivity per partnership transmissibility per year | | |  |  |
|  | Acute stage | | 0.030–0·61 | [7]; Model calibration |
|  | Chronic stage | | 0.027–0·21 | [7]; Model calibration |
|  | AIDS stage | | 0.008–0·27 | [7]; Model calibration |
| Proportion MSM in sexual risk groups | | |  |  |
|  | Highest | | 11% (11-12% IQR) | Model calibration, the sum of the three groups was equal to 100% |
|  | 2^nd^ highest | | 14% (12-16% IQR) |  |
|  | 3^rd^ highest | | 19% (14-23% IQR) |  |
|  | Lowest | | 56% (49-61% IQR) |  |
| Proportion MSM in sexual risk groups | | |  |  |
|  | | Highest | 43.88 (34.95-55.27 IQR) | Model calibration |
|  | | 2^nd^ highest | 9.78 (7.99-12.05 IQR) |  |
|  | | 3^rd^ highest | 2.65 (1.85-3.67 IQR) |  |
|  | | Lowest | 0.20 (0.14-0.29 IQR) |  |
| Mortality rates per year | | |  |  |
|  | Population | | 0.0155 | [8] |
|  | Chronic HIV stage | | 0.114 | [8] |
|  | AIDS stage | | 0.648 | [8] |
|  | On treatment | | 0.0184 | [8] |
| PrEP effectiveness | | | 85% | [9] |
| PrEP regimens discontinuation rate | | | 0.62/year (0.48-0.79 IQR) | Model calibration |
| Primary cost parameters (costs listed are in 2022 euros) from a payer’s perspective | | | | |
| Yearly cost of PrEP^b^ | | | €788.19 | [10]; Local data |
| Yearly cost of ART by diagnosed stages^c^ | | |  |  |
|  | | Timely presenter | €12465.92 | [10, 11] |
|  | | Late presenter | €14541.90 | [10, 11] |
|  | | Advanced presenter | €24372.57 | [10, 11] |

^Note: Abbreviations: AIDS: acquired immunodeficiency syndrome; HIV: human immunodeficiency virus; IQR: interquartile range; PrEP: pre-exposure prophylaxis.^

^a All ranges were uniformly distributed.^

^b Details see Supplementary Table S3^

^c Details see Supplementary Table S4^

### Simulated scenarios

Figure S1. Scenarios simulated for expanding PrEP provision to cover a) 3000 MSM who are currently on the waiting list, and b) 19500 MSM who are PrEP eligible and express intention for PrEP use with formal PrEP provision by the public health services, the Netherlands, 2021-2030.

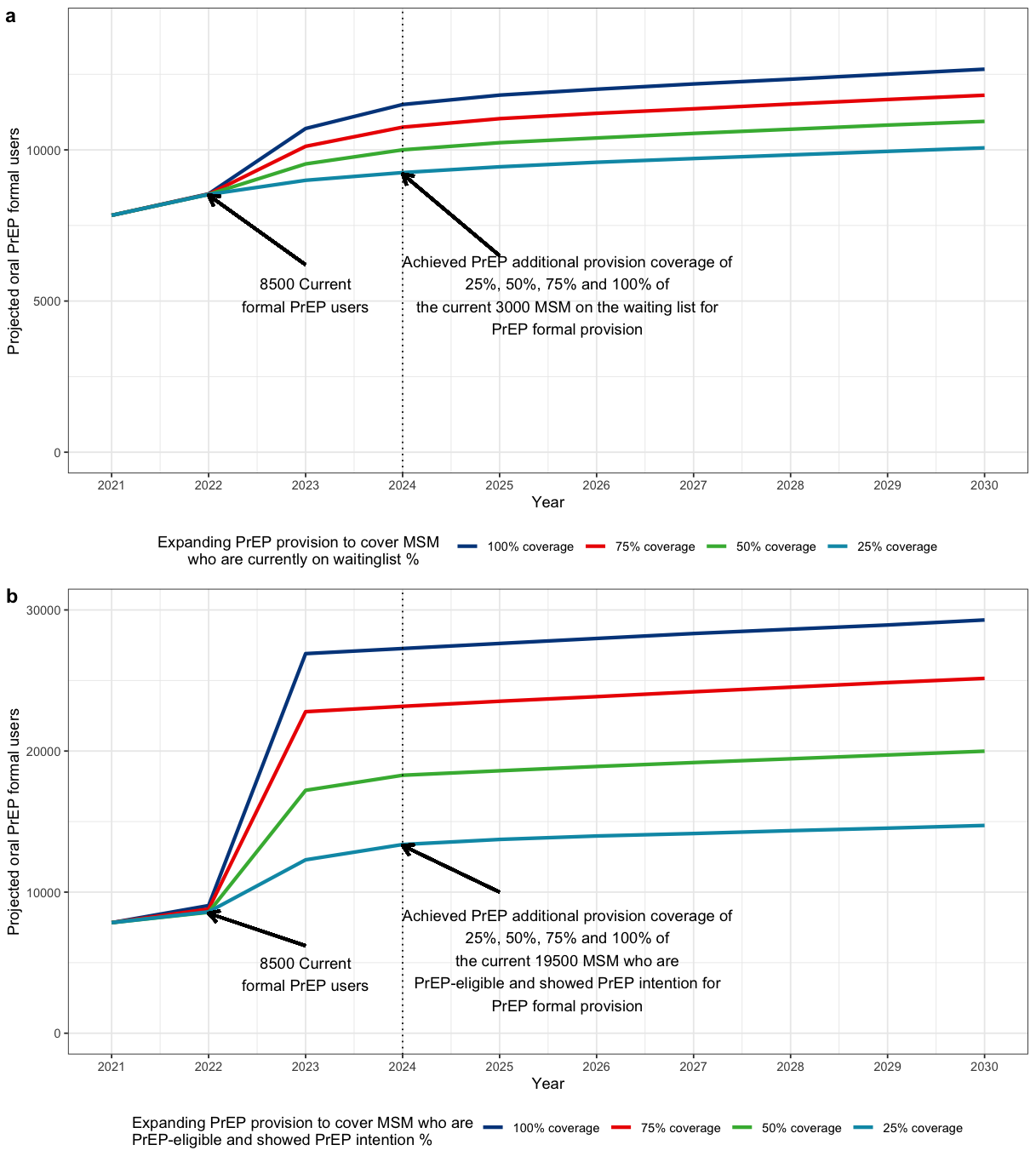

### Utility weighting assumptions

**Table S3 Assumed utility weighting for Quality-adjusted life year (QALY).**

| Status | Utility weight | Reference |
| --- | --- | --- |
| Not infected with HIV/using PrEP | 1 | **[12]** |
| CD4 > 350 cells/ | 0.94 | **[12]** |
| CD4 cell count 200-350 cells/ | 0.82 | **[12]** |
| AIDS stage | 0.7 | **[12]** |
| HIV infected using antiretroviral drug treatment | 0.94 | **[12]** |

### Utility costs

The costs that we included for PrEP were the local price of Tenofovir disoproxil fumarate/emtricitabine (TDF/FTC), the costs of visiting a physician who prescribes PrEP and the costs of monitoring side effects of tenofovir (e.g. creatinine and Urine culture) and screening sexually transmitted infections (STIs, including HIV, syphilis, hepatitis C and other bacterial STIs), according to the Dutch practice guideline [3]. We used unit prices from the year 2022 and recommended unit prices from before the year 2022 were indexed based on annual Dutch inflation as stated by the Central Bureau for Statistics. The annual costs for providing PrEP, including monitoring and costs of the drugs, are €788.19. More detailed information on the included costs of PrEP can be found in Table S3.

**Table S4 Annual costs per service for the use of oral pre-exposure prophylaxis (PrEP) in the Netherlands**

| **Cost scenario** | **Type of service** | **Frequencies** | **Initial visit** | **Annual cost** |
| --- | --- | --- | --- | --- |
| PrEP | HIV screening | Initial visit, 1 month after initiation, after that every 3 months | € 12,95 | € 51,80 |
|  | Hepatitis C | Initial visit, after that every 12 months | € 12,65 | € 12,65 |
|  | Syphilis | Initial visit, after that every 3 months | € 12,95 | € 51,80 |
|  | Chlamydia, Gonorrea | Initial visit, after that every 3 months | € 70,44 | € 281,76 |
|  | HBV screening | Initial visit | € 24,78 | € 0,00 |
|  | Creatinine | Initial visit, 1 month after initiation, after that every 6 months | € 1,60 | € 1,60 |
|  | Urine culture | Initial visit, 1 month after initiation, after that every 12 months | € 28,58 | € 28,58 |
|  | PrEP | Daily intake | - | € 360,00 |
| Total cost PrEP | | |  | € 788,19 |

For the cost of treatments, we took into account the local costs by diagnosed stages, which included timely presenter (CD4<350 cells/mm^3), late presenter (CD4<350 cells/mm^3) and advanced presenter (CD4<200 cells/mm^3), which consist of the costs for antiretroviral drugs and direct healthcare costs, using local data [10, 11]. Direct healthcare costs included outpatient visits to an HIV specialist and to other medical specialists, hospitalization, viral load measurements, CD4 Cell count measurements, and potential additional co-medication for the late and advanced presenter [11]. The annual costs for HIV treatment, including monitoring and costs of the drugs, are €12465.92 for the timely presenter, €14541.90 for the late presenter and €24372.57 for the advanced presenter. More detailed information on the included costs related to treatment can be found in Table S4.

**Table S5 Annual costs per service for the antiretroviral drugs and other direct healthcare costs by diagnosed stage in the Netherlands.**

| **Cost scenario** | **Type of service** | **Timely presenter** | **Late presenter** | **Advanced presenter** | **Reference** |
| --- | --- | --- | --- | --- | --- |
| Treatment | Outpatient visits | 567,6 | 696,6 | 761,1 | [10, 11] |
|  | Inpatient visits | 258,02 | 1222,2 | 7115,92 | [10, 11] |
|  | Viral loads | 437,4 | 486 | 522,45 | [10, 11] |
|  | CD4 Cell count | 318,6 | 413 | 1203,6 | [10, 11] |
|  | co-medication | 0 | 839,8 | 3885,2 | [10, 11] |
|  | ART (TAF/FTC/BIC) | 10884,3 | 10884,3 | 10884,3 | [10, 11] |
| Total treatment | | 12465,92 | 14541,90 | 24372,57 | [10, 11] |

### One-way Sensitivity Analysis for the Costs

**Figure S2. One-way sensitivity analyses of the incremental cost-eﬀectiveness of improving PrEP provision to cover 75% of PrEP-eligible/intending MSM compared with the current PrEP provision in the Netherlands over 40 years, 2022-2062**

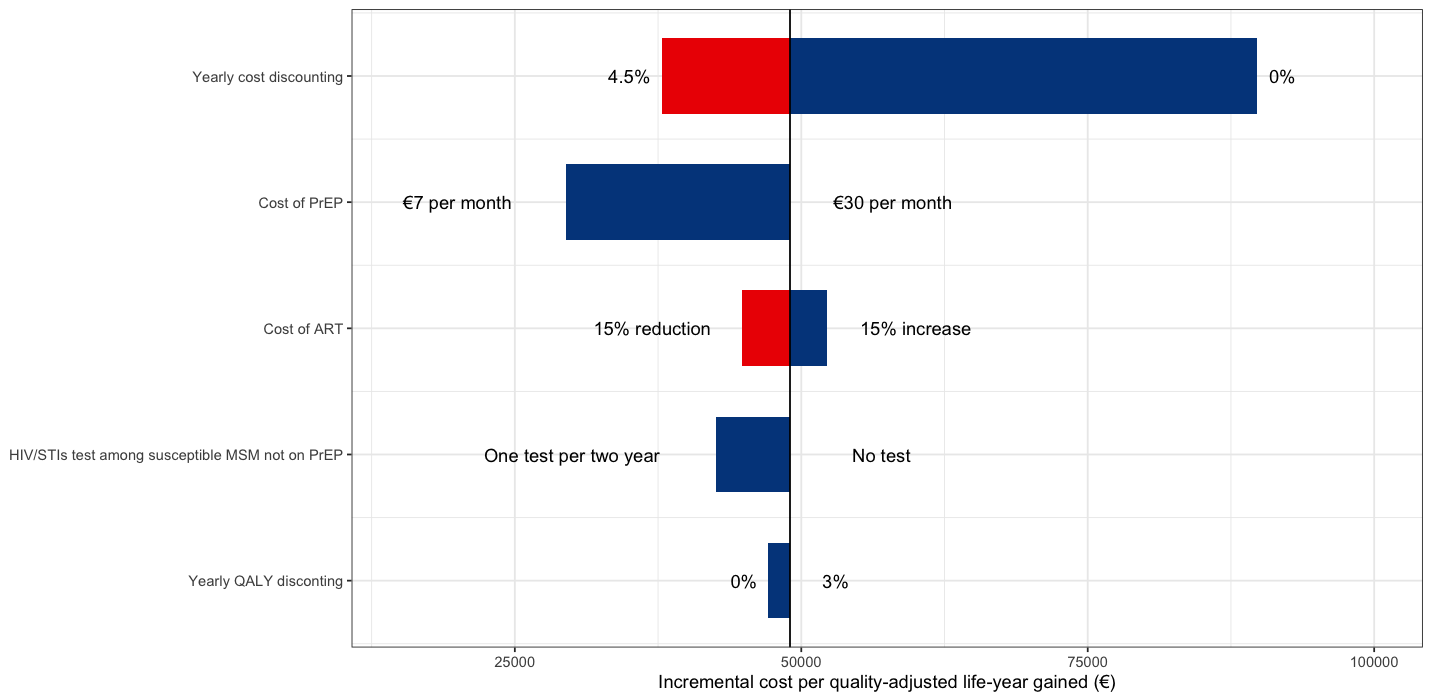
