## Supplemental File S2 for "Epidemiological impact and cost-effectiveness of expanding formal PrEP provision to PrEP-eligible MSM expressing PrEP-intention in the Netherlands"

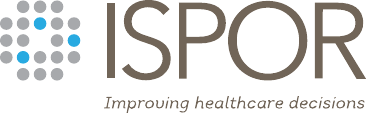

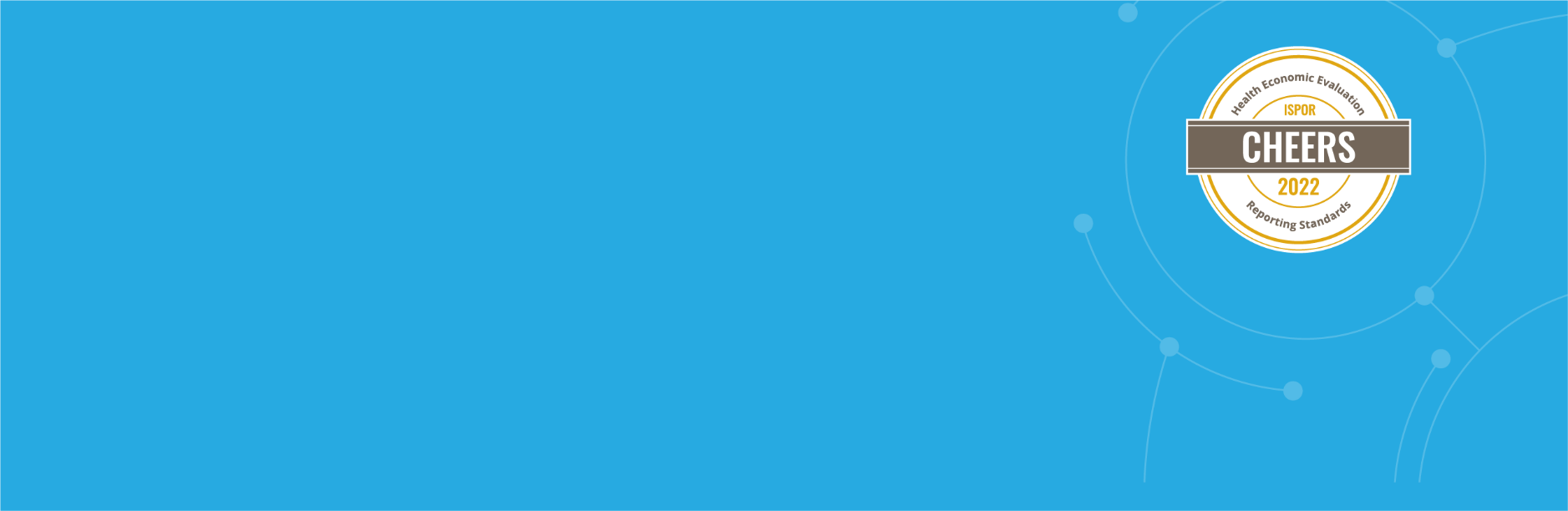

**Consolidated Health Economic Evaluation Reporting Standards
(CHEERS) 2022 Checklist**

The CHEERS 2022 statement replaces the 2013 CHEERS statement, which should no longer
be used. The CHEERS 2022 checklist contains 28 items with accompanying descriptions.
Checklist users should indicate the section of the manuscript where relevant information
can be found. The authors recommend using a section heading with a paragraph number.
If an item does not apply to a particular economic evaluation, checklist users are encouraged
to report “Not Applicable.” If information is otherwise not reported, checklist users are encouraged to

write, “Not Reported.” Users should avoid the term “Not Conducted” as CHEERS is intended to guide and

capture reporting. Additional information on CHEERS 2022 can be found here.

**Title**

**1. Title**

Identify the study as an economic evaluation and specify the interventions being compared.

Epidemiological impact and cost-effectiveness of expanding formal PrEP provision to PrEP-eligible MSM expressing PrEP-intention in the Netherlands

**Abstract**

**2. Abstract**

Provide a structured summary that highlights context, key methods, results, and alternative analyses.

Abstract: page 1

**Introduction**

**3. Introduction: Background and Objectives**

Give the context for the study, the study question, and its practical relevance for decision making in policy or practice.

Introduction

**Methods**

**4. Health economic analysis plan**

Indicate whether a health economic analysis plan was developed and where available.

Supplement S2: Before starting the study, a Health Economics Analysis Plan was established by a team of health economists (SP, DvdV).

**5. Study population**

Describe characteristics of the study population (such as age range, demographics, socioeconomic, or clinical characteristics).

Estimating the number of PrEP-eligible/intending MSM

**6. Setting and location**

Provide relevant contextual information that may influence findings.

Introduction

**7. Comparators**

Describe the interventions or strategies being compared and why chosen.

Mathematical modelling of expanding formal PrEP provision scenarios

**8. Perspective**

State the perspective(s) adopted by the study and why chosen.

Cost-effectiveness of expanding formal PrEP provision

**9. Time horizon**

State the time horizon for the study and why appropriate.

Cost-effectiveness of expanding formal PrEP provision

**10. Discount rate**

Report the discount rate(s) and reason chosen.

Cost-effectiveness of expanding formal PrEP provision

**11. Selection of outcomes**

Describe what outcomes were used as the measure(s) of benefit(s) and harm(s).

Cost-effectiveness of expanding formal PrEP provision

**12. Measurement of outcomes**

Describe how outcomes used to capture benefit(s) and harm(s) were measured.

Cost-effectiveness of expanding formal PrEP provision

**13. Valuation of outcomes**

Describe the population and methods used to measure and value outcomes.

Cost-effectiveness of expanding formal PrEP provision

**14. Measurement and valuation of resources and costs**

Describe how costs were valued.

Cost-effectiveness of expanding formal PrEP provision and Supplementary S2 File

**15. Currency, price date, and conversion**

Report the dates of the estimated resource quantities and unit costs, plus the currency and year of conversion.

Supplementary S2 File

**16. Rationale and description of model**

If modeling is used, describe in detail and why used. Report if the model is publicly available and where it can be accessed.

Mathematical modelling of expanding formal PrEP provision scenarios

**17. Analytics and assumptions**

Describe any methods for analyzing or statistically transforming data, any extrapolation methods, and approaches for validating any model used.

Not applicable

**18. Characterizing heterogeneity**

Describe any methods used for estimating how the results of the study vary for subgroups.

Supplementary S2-S3 File: Cost-effectiveness analysis and One way sensitivity analysis

**19. Characterizing distributional effects**

Describe how impacts are distributed across different individuals or adjustments made to reflect priority populations.

Not applicable

**20. Characterizing uncertainty**

Describe methods to characterize any sources of uncertainty in the analysis.

Supplementary S3 File: One way sensitivity analysis

**21. Approach to engagement with patients and others affected by the study**

Describe any approaches to engage patients or service recipients, the general public, communities, or stakeholders (eg, clinicians or payers) in the design of the study.

Before starting the study, a Health Economics Analysis Plan was established by a team of health economists (SP, DvdV)

**Results**

**22. Study parameters**

Report all analytic inputs (eg, values, ranges, references) including uncertainty or distributional assumptions.

Supplement S1 File

**23. Summary of main results**

Report the mean values for the main categories of costs and outcomes of interest and summarize them in the most appropriate overall measure.

Cost-effectiveness of expanding formal PrEP provision and Supplementary S2-S3 File

**24. Effect of uncertainty**

Describe how uncertainty about analytic judgments, inputs, or projections affects findings. Report the effect of choice of discount rate and time horizon, if applicable.

Supplementary S3 File

**25. Effect of engagement with patients and others affected by the study**

Report on any difference patient/service recipient, general public, community, or stakeholder involvement made to the approach or findings of the study.

Not applicable

**Discussion**

**26. Study findings, limitations, generalizability, and current knowledge**

Report key findings, limitations, ethical, or equity considerations not captured and how these could impact patients, policy, or practice.

Discussion section

**Other Relevant Information**

**27. Source of funding**

Describe how the study was funded and any role of the funder in the identification, design, conduct, and reporting of the analysis.

Funding/support statement

**28. Conflicts of interest**

Report authors’ conflicts of interest according to journal or International Committee of Medical Journal Editors requirements.

Conflicts of Interest statement
